# Recruitment location influences bias and uncertainty in SARS-CoV-2 seroprevalence estimates

**DOI:** 10.1101/2021.02.03.21251011

**Authors:** Tyler S. Brown, Pablo Martinez de Salazar Munoz, Abhishek Bhatia, Bridget Bunda, Ellen K. Williams, David Bor, James S. Miller, Amir M. Mohareb, Julia Thierauf, Wenxin Yang, Julian Villalba, Vivek Naranbai, Wilfredo Garcia Beltran, Tyler E. Miller, Doug Kress, Kristen Stelljes, Keith Johnson, Daniel B. Larremore, Jochen Lennerz, A. John Iafrate, Satchit Balsari, Caroline O. Buckee, Yonatan H. Grad

## Abstract

The initial phase of the COVID-19 pandemic in the US was marked by limited diagnostic testing, resulting in the need for seroprevalence studies to estimate cumulative incidence and define epidemic dynamics. In lieu of systematic representational surveillance, venue-based sampling was often used to rapidly estimate a community’s seroprevalence. However, biases and uncertainty due to site selection and use of convenience samples are poorly understood. Using data from a SARS-CoV-2 serosurveillance study we performed in Somerville, Massachusetts, we found that the uncertainty in seroprevalence estimates depends on how well sampling intensity matches the known or expected geographic distribution of seropositive individuals in the study area. We use GPS-estimated foot traffic to measure and account for these sources of bias. Our results demonstrated that study-site selection informed by mobility patterns can markedly improve seroprevalence estimates. Such data should be used in the design and interpretation of venue-based serosurveillance studies.

## Introduction

Studies estimating SARS-CoV-2 seroprevalence have been critical to our understanding of COVID-19 pandemic dynamics, particularly in times when diagnostic testing was limited and the extent of community spread was unknown [1–4]. However, many of these estimates were derived from non-representational convenience sampling [4–6], thus making these estimates subject to multiple sources of bias and uncertainty that remain poorly understood [7]. Low precision can obscure differences in estimated seroprevalence between populations, limiting efforts to understand heterogeneity in epidemic intensity or vaccination coverage and reducing the effectiveness of public health interventions. As cost, speed, and logistical concerns continue to motivate the use of convenience sampling [8], identifying and accounting for bias and uncertainty are therefore critical to improving the design and interpretability of these studies.

Convenience sampling involves inherently non-uniform sampling across demographic or geographic subgroups. For example, in venue-based (or “walk-up”) studies, in which participants are recruited from among visitors to a central or highly trafficked location [4,9], the geographic distribution of participants is expected to be skewed toward individuals living closer to the study location. A similar concern applies to studies using discarded blood samples, where the catchment area of a given hospital or clinical laboratory may strongly constrain the geographic distribution of samples available for analysis [5].

Multiple recent studies have underscored the utility of GPS- and mobile phone-associated human mobility data in understanding COVID-19 epidemiology. These studies have clarified the role of highly-visited “super-spreader” locations [10,11] in local COVID-19 epidemics and identified higher aggregate mobility as a predictor of neighborhood-level COVID-19 risk [3]. Prospective applications of these data sources, including their use in the design and implementation of epidemiological studies, are limited. Here, we examine seroprevalence estimates obtained via venue-based sampling [4,9], with direct applications to other forms of convenience sampling [5,12,13]. We performed a venue-based serosurveillance study in Somerville, Massachusetts and used these data to analyze bias and uncertainty arising from inherent geographic variation in sampling intensity. This analysis showed that substantial loss of precision can occur if the distribution of sampling intensity poorly matches the geographic distribution of true seropositive individuals in the study area. As GPS-estimated foot traffic offers a proxy measurement for the geographic distribution of visitors to different locations, we evaluated the extent to which informed selection of locations for venue sampling reduces uncertainty and bias introduced by geographic heterogeneity in underlying seropositivity. Our results thus offer an approach to significantly improve the design and interpretation of seroprevalence studies that use venue-based convenience sampling with little if any impact on cost and speed.

## Methods

### SARS-CoV-2 seroprevalence study design and participant information

We obtained serological and participant demographic data for 398 asymptomatic adults tested at a temporary study site near an essential business location in Somerville, Massachusetts. The study was conducted over 4 days (June 4^th^, 5^th^, 8^th^, and 9^th^, 2020), approximately 6 weeks after the first wave of the COVID-19 epidemic peaked in Massachusetts [14]. The study was designated minimal risk human subjects research and approved by institutional review boards at Massachusetts General Hospital and the Harvard T.H. Chan School of Public Health (Protocol number: 2020P001081). The study recorded participant demographic information including age, gender, and self-reported home locations (by ZIP code and electoral ward). We also collected information on how participants learned about the study in order to distinguish participants directly recruited on site at the study location from those who learned about the study from friends, family, or social media. We did not advertise or announce enrollment for the study prior to its implementation, with the goal of increasing the proportion of individuals recruited on site at the study location. We used this data to calculate 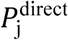, the proportion of all directly recruited participants from Somerville with self-reported home locations in electoral ward *j* ∈ {1,…,7}. We refer to 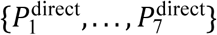 as the “survey participant catchment distribution.”

### Public health acute infection data

We obtained data on 916 PCR-confirmed COVID-19 cases with documented home addresses in Somerville (collected from the onset of the epidemic through June, 2020) from the Massachusetts Virtual Epidemiologic Network (MAVEN). COVID-19 cases are reported to MAVEN by state and local health agencies, and cases are designated as “confirmed” if they have a positive result for SARS-CoV-2 RNA detection using an FDA-approved molecular amplification detection test, for example RT-PCR. The total number of Somerville residents tested for SARS-COV-2 via RT-PCR was also obtained from MAVEN. Data were anonymized and aggregated by electoral ward prior to analysis. We calculated the cumulative incidence of PCR-confirmed infections by ward 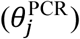 and the proportion of all PCR tests with positive results (“PCR positivity”).

### GPS-estimated business foot traffic

We used GPS-estimated foot traffic (SafeGraph, *safegraph*.*com*) to approximate the distribution of home locations for daytime visitors for different locations of interest, which we refer to as the “GPS-estimated catchment area.” We use these data to estimate home and work locations at the level of census block group (CBG) for visitors to designated points-of-interest, such as businesses, and specific CBGs. We used CBG-level visitor data for June 2020 as the primary data source for visitors in our analysis.

These data were filtered for CBGs with low visitor counts and re-aggregated from CBGs to electoral wards prior to analysis (Supplementary Material, Figures S1 and S2). We used the filtered, re-aggregated data to obtain GPS-estimated visitor catchment distributions for two locations: (1) the actual study venue, located in Somerville electoral Ward 2 (denoted *V*_j_^site^) and (2) a hypothetical alternative study venue located in Somerville electoral Ward 1 (denoted *V*_j_^alt^).

### Simulations

We used numerical simulation to examine bias and uncertainty in estimated seroprevalence 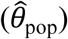 as a function of subgroup sizes, true subgroup seropositivity, and sample allocation. Here, “subgroup” is used to describe any potential stratification of the overall population, including by demographic and/or geographic characteristics. Briefly, we used demographic [15] and SARS-CoV-2 acute infection data from Somerville, MA to generate a simulated population with varying true seropositivity *θ*_j,k_ across subgroups stratified by age group *j* and location *k* (where locations are electoral wards in Somerville). Briefly, the simulation randomly draws *n*_j,k_ individuals from each subgroup, calculates weighted population-level seroprevalence (adjusted for serological test performance), and repeats this process 10000 times to generate distributions of 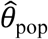 values. We report *W*, the width of the 95th percentile interval for each distribution, as an approximate measure of uncertainty for 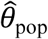 for a given set of simulation parameters. Additional details are available in the Supplementary Material. *R* code for the numerical simulations is available at *https://github.com/svsero/COVID19serosurveillance-Somerville*.

## Results

### Study participant catchment distributions and geographic heterogeneity in COVID-19 epidemic intensity

We first examined how participant catchment distributions align with, or mismatch, the geographic distribution of seropositive individuals in a given study area. We observed that the survey participant catchment distribution in our serosurveillance study was skewed strongly toward locations near the study site (Figure 1A). Among directly recruited participants with home locations in Somerville, 43% (43/100) reported home locations in Somerville Ward 2 (where the study site was located) compared to 4% in Ward 1 and 4% in Ward 4. In contrast, the cumulative incidence of PCR-confirmed SARS-CoV-2 infections 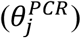 was approximately three-fold higher in electoral Ward 1 compared to Wards 2 and 6 (Figure 1B) and the proportion of SARS-CoV-2 PCR tests with positive results was approximately five-fold higher (Supplementary Figure S3A). Both of these proxy measures of epidemic intensity (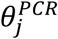 and PCR test positivity) are limited by potential biases, some of which are likely to still be present even if PCR testing rates are relatively equal by ward [16]. Nonetheless, these measures suggest substantial heterogeneity in the underlying epidemic intensity, with an apparent higher rate of previously infected individuals (as a proportion of the population) in Somerville Wards 1 and 4. Thus, we observed that the venue location chosen for this study, and its associated survey participant catchment distribution, resulted in relative under-sampling of wards with expected higher seropositivity and oversampling of those with lower expected seropositivity (Figure 1C).

**Figure 1.**
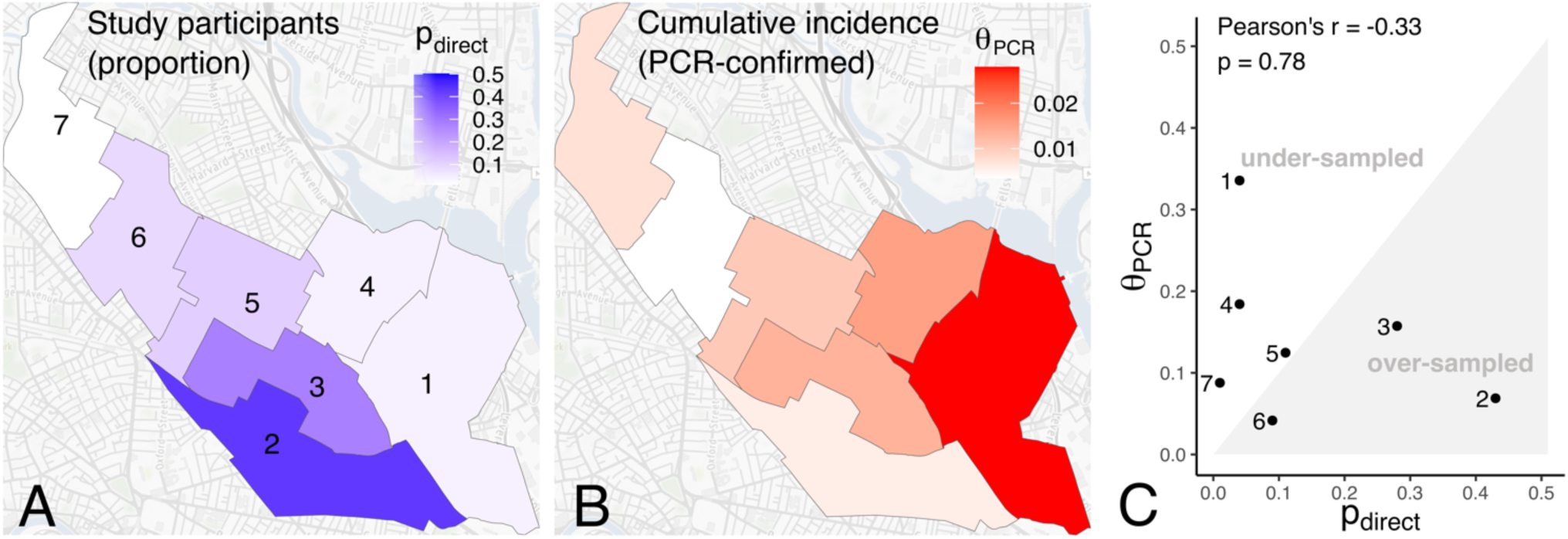
Sample allocation and geographic heterogeneity in proxy measures of epidemic intensity. (A) Survey participant catchment distribution. Wards are shaded by 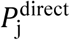, the proportion of all directly recruited participants from each of Somerville Wards 1-7; (B) Cumulative incidence of prior PCR-confirmed SARS-CoV-2 infections by Ward as of June 8th, 2020 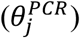; (C) Correlation between 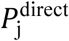 and 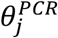. Significance of the correlation is calculated via permutation testing, as described in the Supplementary Material.

### GPS-estimated visitor catchment distributions

Recognizing that survey participant catchment distributions can be poorly matched to the underlying geographic distribution of seropositivity, we explored the use of GPS-estimated foot traffic data as a tool for evaluating actual or candidate locations for venue-based sampling. We evaluated correlations between the observed participant catchment distribution for the actual Somerville study location, GPS-estimated visitor catchment distributions for this site and a hypothetical alternative site in Somerville Ward 1, and the cumulative incidence of PCR-confirmed infections by ward. The participant catchment distribution at the actual study site 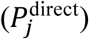 closely matched its corresponding GPS-estimated visitor catchment distribution, *V*_j_^*site*^(Pearson’s *r*= 0.90, *p*=0.0131, Figure S4). However, the GPS-estimated visitor catchment distribution for the actual study site was poorly correlated with the cumulative incidence of PCR-confirmed infections (*r*= -0.11, *p*= 0.55, Figure 2C).

**Figure 2.**
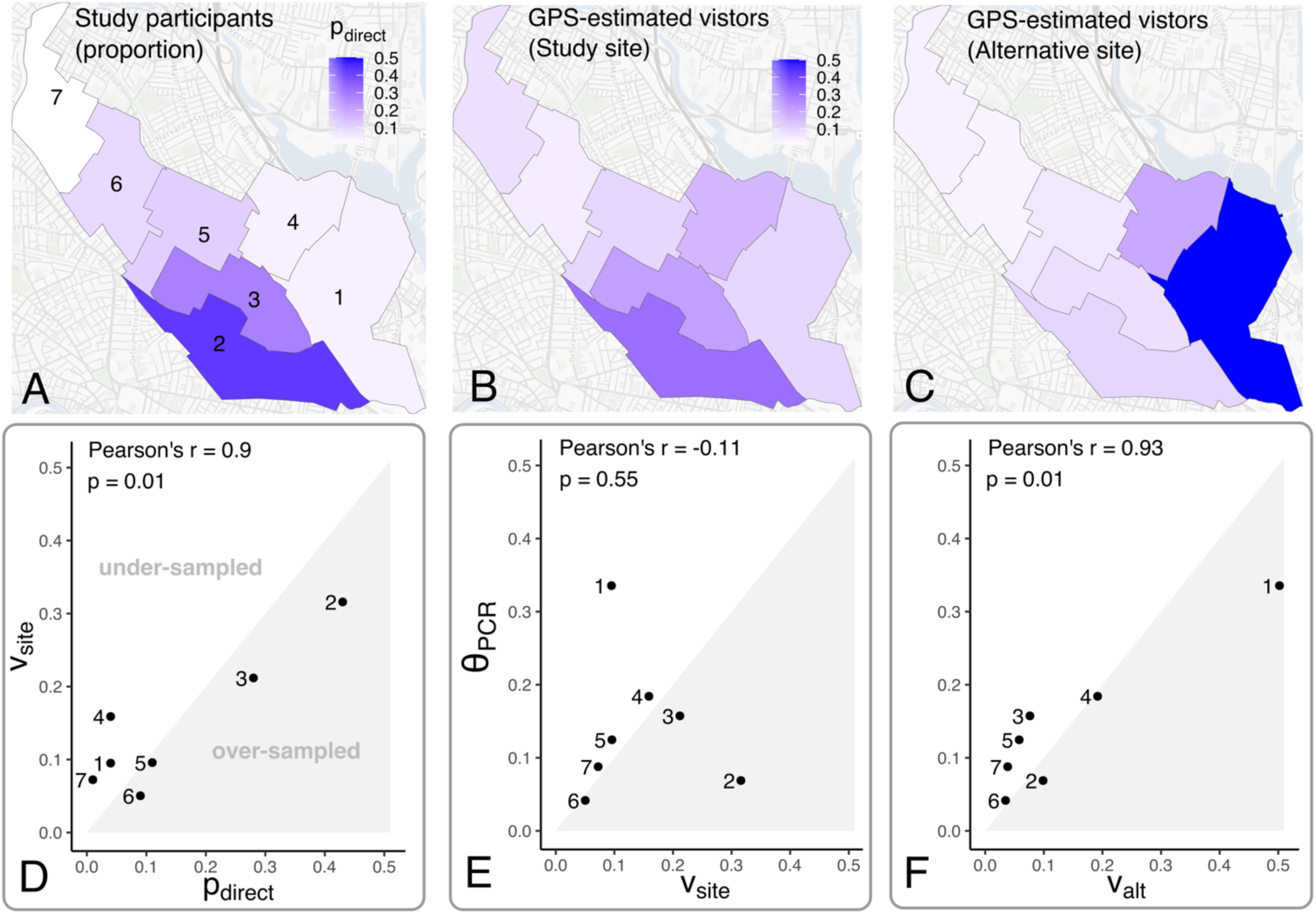
GPS-estimated visitor catchment distributions for actual and hypothetical alternative study sites. GPS-estimated visitor catchment distributions for (A) the actual study location *V*_j_^site^ or (B) a hypothetical alternative study site in Somerville Ward 1 *V*_j_^alt^. Correlations between *V*_j_^site^, *V*_j_^alt^, and the cumulative incidence of PCR-confirmed SARS-CoV-2 infection 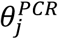 are shown in (C) and (D).

We evaluated whether choosing an alternative study site could improve the correlation between sample allocation and cumulative incidence of PCR-confirmed infections by ward, with the goal of reducing uncertainty in estimated seroprevalence. We observed that the GPS-estimated visitor catchment distribution at the alternative site (*V*_j_^*alt*^) is strongly correlated with ward-level cumulative incidence of PCR-confirmed infections (*r*=0.93, *p*=0.0072, Figure 2D). If subgroup sizes are known and differences in subgroup-level seropositivity can be inferred or assumed, allocating more samples to larger subgroups and those with higher expected seropositivity will improve precision for weighted population-level seroprevalence estimates (Figure S5 and Equation 2) [17]. This suggests that, if the GPS-estimated visitor catchment distribution reliably predicts the survey participant catchment at the alternative study site, this location would yield improved sample allocation across geographic subgroups that more closely approximates optimal sample allocation.

### Venue location and uncertainty in SARS-CoV-2 seroprevalence estimates

We used numerical simulation to quantify uncertainty in estimated SARS-CoV-2 seroprevalence under different survey participant catchment distributions. Specifically, we constructed a simple synthetic population with seven geographic subgroups, corresponding to the seven electoral wards in Somerville, each divided into three age-based subgroups. We specified the size of each subgroup using local census data [15] and specified the true underlying seropositivity for each age-location subgroup by assuming these values are proportional to the observed cumulative incidence of PCR-confirmed infections for each subgroup. Using this model, we compared three sample allocation scenarios: (1) optimal allocation, in which the number of individuals sampled from each age-location subgroup is specified to optimally reduce uncertainty in the resulting seroprevalence estimates (per Equation 2 in the Supplementary Information); (2) allocation according to the observed survey participant catchment distribution for the actual study site; (3) allocation according to the GPS-estimated visitor distribution at the hypothetical alternative study site. (Additional details on model specification and sensitivity testing for model parameters are available in the Supplementary Information.)

We observed 1.5- to 2-fold higher uncertainty when sampling effort was allocated according to the participant catchment distribution at the study site compared to the alternative site or optimal allocation (Figure 3). This observation suggests that choice of recruitment location can result in suboptimal sample allocation and higher uncertainty.

**Figure 3.**
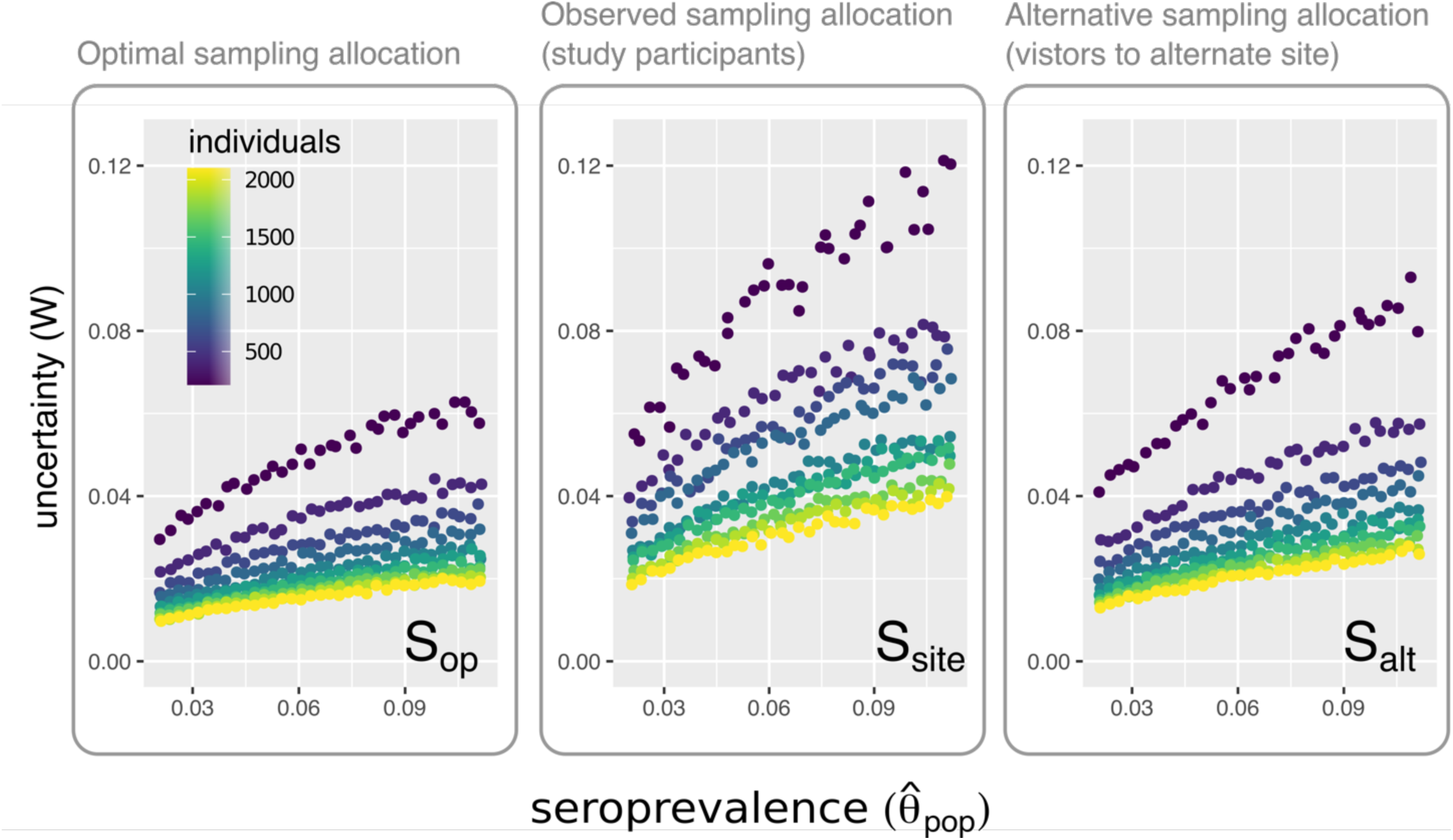
Uncertainty in estimated SARS-CoV-2 seroprevalence obtained using different sample allocation strategies. The uncertainty (*W*, the width of the 95th percentile interval for 10000 estimated seroprevalence values) versus mean estimated seroprevalence for different values of *n* (the total number of individuals sampled) when individuals are sampled using (1) in the left panel, the optimal sample allocation according to Equation 2 in the Supplementary Material (***S***_***op***_); (2), in the center panel, the sampling distribution of participants in the Somerville seroprevalence survey (***S***_***site***_); (3) the sampling distribution at the proposed alternative study site (***S***_**alt**_).

### Bias due to unappreciated heterogeneity in seropositivity across geographic subgroups

Biased seroprevalence estimates can result if geographic heterogeneity in sample allocation results in substantial over- or under-sampling in locations with higher (or lower) seropositivity, and if procedures for generating weighted prevalence estimates do not appropriately account for geographic heterogeneity in underlying seropositivity. We compared estimated seroprevalence versus true seroprevalence for numerical simulations in which the final seroprevalence estimates were weighted by the sampling probability for each age-location group or by the sampling probability of each age subgroup alone (Figure 4). The first weighting procedure accounts for heterogeneity across age and location subgroups, whereas the second procedure accounts only for heterogeneity across age subgroups. Using the second procedure resulted in over- or under-estimation of seroprevalence, depending on whether sample allocation enriches for participants from areas with high or low underlying seropositivity, respectively.

**Figure 4.**
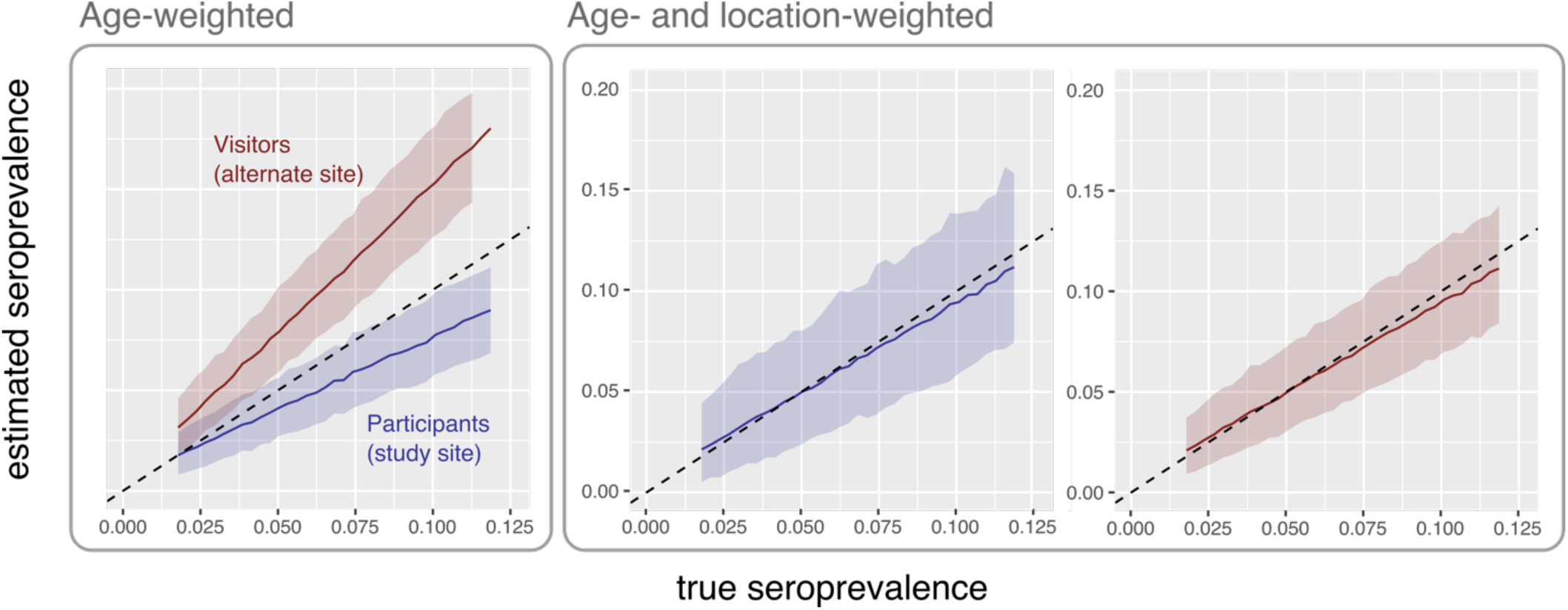
Bias in estimated seroprevalence by sampling strategy and weighting procedures. Left: Estimated seroprevalence, weighted only by age subgroups. Right: Estimates weighted by age-location subgroups are shown in the right panel. Blue: Sample allocation specified by the observed participant distribution catchment distribution in the Somerville study. Red: Sample allocation specified by the catchment distribution of GPS-estimated visitors to the proposed alternate study site. Dotted line indicates where estimated equals true seroprevalence.

## Discussion

Convenience sampling, despite its inherent limitations, may have continued utility in the public health response to the COVID-19 pandemic and for infectious disease outbreaks generally. Cost and logistical considerations may limit the feasibility of randomized structured sampling, particularly in resource-constrained contexts or in situations where census data, population rosters, or household mapping data are unavailable or unreliable. Certain forms of convenience sampling are better suited for reaching important subgroups compared to structured approaches. Lower-wage or frontline workers who are at higher risk of SARS-CoV-2 exposure [18-20], including undocumented workers [20], may be less likely to participate if recruited using conventional survey outreach methods (e.g., mail or phone contact) due to constraints on their time [21-23] and lack of incentives [21]. Convenience sampling at highly visited community locations such as essential businesses may be an attractive alternative to structured sampling in this important population, similar to sampling approaches developed to study so-called “hidden populations” [24].

Geographic heterogeneity in SARS-CoV-2 epidemic intensity within cities has been a repeatedly observed feature of the pandemic [1–3]. This phenomenon poses unique challenges for seroprevalence studies that employ venue-based and other convenience sampling strategies, in which sample allocation across subgroups cannot be pre-specified and is non-uniform across geographic space.

Examining data from our seroprevalence study in Somerville, MA, we observed that venue-based sampling resulted in substantial undersampling of areas where proxy measures (cumulative incidence of PCR-confirmed infections and PCR test positivity rates) suggest higher epidemic intensity. This mismatch between the survey participant catchment distribution and the geographic distribution of seropositive individuals can result in suboptimal sample allocation and higher variance in resultant seroprevalence estimates.

Study locations can have widely divergent participant catchment distributions. These distributions can be heavily enriched for participants living in the immediate vicinity of the study location. This limitation has important implications for bias and uncertainty of resulting seroprevalence estimates (as detailed here) and raises questions about potential undersampling or exclusion of important subgroups in venue-based studies. Multiple studies have identified geographic location as a strong surrogate for multiple risk factors associated with severe infection, hospitalization, and/or death due to COVID-19 [25] and undersampling in neighborhoods where these risk factors co-localize together can compromise the reliability and interpretability of seroprevalence estimates. Recruiting participants directly from such communities, where rates of COVID-19 related hospitalization and deaths are often higher, has yielded seroprevalence estimates that are substantially higher than city-level or state-level estimates [4,9]. Notably, the areas that were most undersampled in our study strongly overlap neighborhoods with lower socioeconomic status, larger proportions of non-white residents, lower proportions of English-speaking households (Figure 1A, Supplementary Figure S3C).

Our work has three practical findings that are applicable to the design, implementation, and interpretation of convenience-based seroprevalence studies.

1. Uncertainty in population-level seroprevalence estimates is minimized when sample allocation is proportional to the size and underlying seropositivity of individual subgroups in the population (Equation 2 and Figure S5). The practical application of this finding may be limited because of challenges in reliably ascertaining differences in the underlying seroprevalence between subgroups *a priori*. For example, this may arise when access to diagnostic testing for acute infections is limited or disparate across subgroups. However, even in this situation, allocating sampling effort proportional to subgroup sizes alone can substantially reduce uncertainty (Figure S1). Applying this finding may also be difficult because an inherent feature of venue-based sampling is that allocation of sampling effort is not pre-specified, but instead results from a stochastic process that depends on the location of the study venue. Thus, although optimal sample allocation is likely not achievable via venue-based sampling, careful selection of venue locations, with the objective of enriching for participants from geographic subgroups with larger populations and/or higher expected seroprevalence, can at least improve sample allocation and help reduce uncertainty.
2. GPS-estimated foot traffic can inform the selection of venue-based recruitment locations. The GPS-estimated visitor catchment distribution at our study location correlated closely with the survey participant catchment distribution. Validation against other data sources that directly measure the geographic distributions of visitors to locations of interest (for example, aggregated geographic and registration data from COVID-19 mobile testing programs) can help further evaluate this potentially important data source.
3. Convenience sampling can produce biased seroprevalence estimates if geographic heterogeneity in underlying subgroup-level seropositivity is not properly accounted for (Figure 5). Consistent with prior studies [2, 3], we observed that COVID-19 incidence can vary widely, even over a relatively small geographic area. To avoid this problem, studies that employ convenience sampling should collect geographic data on participants’ home locations that is granular enough to capture potential geographic heterogeneity in seroprevalence within the study area. This information, combined with data on the catchment distributions of individual recruitment sites (including GPS-estimated foot traffic data, as examined here), can be used to quantify what would otherwise be an unmeasured source of bias in resulting seroprevalence estimates.

Multiple considerations are important for contextualizing our findings and the recommendations above. These include participation bias that may result in exclusion of individuals with disabilities or others who are less likely to leave their homes. Methods designed to account for participation bias, including those developed for use with time-location sampling [26], may be applicable here. Likewise, collecting information on non-respondents in venue-based sampling—for example, brief demographic surveys collected before recruitment for serological testing—can help measure and account for potential sources of participation bias.

The GPS-estimated foot traffic data used in our study have several important limitations. Identification of visitors and their home locations in this data may be biased by differences in mobile device usage between demographic groups, potentially under-sampling visitors from important populations or oversampling others. In addition, this data can be sparse and thus more subject to stochastic variation when only small numbers of users are captured in either the point of interest or home location. Different forms of participation bias (described above) are expected to skew the geographic distribution of study participants away from GPS-estimated catchment distributions, potentially making it difficult to use this data source in real-world public health practice.

Lastly, we make several simplifying assumptions in our numerical model. The numerical model assumes that the true seropositivity in each age-location subgroup is proportional to its observed cumulative incidence of PCR-confirmed SARS-CoV-2 infections (per local public health data from Somerville, MA). However, wards with higher PCR positivity rates (an indicator of greater epidemic intensity) have relatively the same rates of overall PCR testing per capita (Supplementary Figure S3B), indicating that there were gaps in testing effort in areas of Somerville with more incident infections overall [16]. The assumed true underlying seroprevalence of each age-location group, which is specified using the observed cumulative incidence of PCR-confirmed infection and does not account for the testing gap described above, are less dispersed across age-location groups than what would be expected if PCR testing effort better matched epidemic intensity by ward (i.e., greater PCR testing effort in heavily impacted areas would likely reveal even larger differences in cumulative incidence between wards). This misspecification, and resultant smaller dispersion in assumed true cumulative incidence by ward, is expected to result in more conservative values for the uncertainty in estimated population-level seroprevalence; otherwise, this limitation is not expected to change our primary findings from the numerical model.

In summary, we have examined how geographic heterogeneity in sample allocation, combined with underlying heterogeneity in geographic distribution of seropositive individuals, can influence seroprevalence estimates derived from venue-based sampling. Our findings are relevant to studies employing venue-based recruitment and are also applicable to other kinds of convenience sampling, for example, studies using a hospital’s discarded blood specimens from patients drawn from the hospital’s geographic catchment area.

## Data Availability

The data used in this study are available either via publicly available sources (for business foot traffic data) or via request of the study authors (serological data).

