## Supplementary Information for "Recruitment location influences bias and uncertainty in SARS-CoV-2 seroprevalence estimates"

#### GPS-estimated foot-traffic data: Filtering and aggregation

We re-aggregated CBG data by electoral ward to match survey data on study participants' home locations using the following procedure: (1) A map of the CBG-level visitor data (where the number of visitors from each CBG is stored as an attribute of its corresponding polygon on the map) was rasterized, using the assumption that visitor counts are uniformly distributed across each polygon; (2) The rasterized polygons were reapportioned to the alternative geometry (wards) using the ``zonal statistics'' function in QGIS (https://qgis.org). For privacy purposes, the third-party data service does not report home location information for CBGs with $< 2$ visitors, and CBGs with $< 4$ visitors are all reported as having exactly 4 visitors. Given the potential uncertainty this introduces for wards with low visitor counts, we filtered the data to exclude wards contributing $< 5$ reported visitors and those representing less than $1\%$ of total visitor traffic to the study site CBG (Supplementary Figures S1 and S2).

#### Optimal sample allocation with heterogeneous seropositivity across subgroups

To examine how sample allocation influences uncertainty in seroprevalence estimates when seropositivity is heterogeneous across subgroups, we modeled differences in subgroup size, subgroup seropositivity, and sample allocation. We examined a simple two subgroup system with total population size of $N = 100000$, $n = 1000$ individuals undergoing serological testing, true population seroprevalence of 5% and 20%. We let $d_1$ and $\pi_1$ both vary across $\{0.1, \ldots, 0.9\}$ (with corresponding values $d_2 = 1 - d_1$ and $\pi_2 = 1 - \pi_1$). For each parameter combination, we report values for the width of the 95% confidence interval ($W$) as a proxy measure of uncertainty, and the proportion of samples allocated to population 1 that minimizes this uncertainty ($\mathit{arg\,min}_{p_1} W(p_1)$).

Figure S3 shows that $\mathit{arg\,min}_{p_1} W(p_1)$ increases with increasing values of $d_1$ and $\pi_1$, i.e. as the size of a given subgroup and its true seropositivity increase, increasing the relative proportion of individuals sampled from this subgroup decreases variance in the resulting population-weighted seroprevalence estimates.

We then generalized these findings to populations with more than two subgroups of interest, i.e. $k$ subgroups indexed by $i \in \{1, \ldots, k\}$. Let $d_i$ be the proportion of the total population in subgroup $i$, let $\theta_i$ be the true underlying seropositivity in subgroup $i$, and let $n_i$ represent the number of individuals who undergo serological testing in each subgroup. Following [1], if the seroprevalences across groups are assumed to be independent of one another, the maximum likelihood estimate of $\theta_i$ is given by

$$\hat{\theta}_i = \frac{n_{i,+}/n_i - u}{1 - u - v}$$

where $n_{i,+}$ is the number positive serologic tests among $n_i$ individuals tested, $u = 1 - \mathit{specificity}$, and $v = 1 - \mathit{sensitivity}$. We assume that sensitivity and specificity of serological testing are uniform across subgroups. The variance for this estimator is

$$\mathrm{Var}\left[\hat{\theta}_i\right] = \frac{\left[u + \hat{\theta}_i(1 - u - v)\right]\left[1 - u - \hat{\theta}_i(1 - u - v)\right]}{n_i(1 - u - v)^2}$$

The variance of the resulting weighted seroprevalence estimate $\hat{\theta}_{\text{pop}} = \sum_i d_i \hat{\theta}_i$ depends on the variances of each $\hat{\theta}_i$.

$$\begin{aligned}\text{Var}\big[\hat{\theta}_{\text{pop}}\big] &\approx \sum_i d_i^2 \,\text{Var}\big[\hat{\theta}_i\big] \\ &\approx \sum_i d_i^2 \frac{\big[u + \hat{\theta}_i(1-u-v)\big]\big[1-u-\hat{\theta}_i(1-u-v)\big]}{n_i(1-u-v)^2} \quad (1)\end{aligned}$$

Finding $\mathbf{n} = (n_1, \ldots, n_K)$ that minimizes $\text{Var}\big[\hat{\theta}_{\text{pop}}\big]$ follows from the derivation of the Neyman allocation using the method of Lagrange multipliers [2,3]. Equation (1) is of the form $f(\mathbf{n}) = \sum_i \frac{c_i}{n_i}$ and is subject to the constraint $\sum_i n_i = n$, where $n$ is the total number of individuals undergoing testing. The Lagrangian formulation for this constrained optimization problem is

$$\nabla f(\mathbf{n}) = \lambda \nabla g(\mathbf{n})$$

$$f(\mathbf{n}) = \sum_i^K \frac{c_i}{n_i}$$

$$g(\mathbf{n}) = \sum_i^K n_i = n$$

which can be solved as

$$\begin{aligned}\frac{\partial f}{\partial n_i} &= \lambda \frac{\partial g}{\partial n_i} \\ -\frac{c_i}{n_i^2} &= \lambda \\ n_i &\propto \sqrt{c_i}\end{aligned}$$

Applying this solution to Equation (3), the value $n_i$ that minimizes the variance of $\hat{\theta}_{\text{pop}}$ is given by

$$n_i \propto d_i \sqrt{[u + \theta_i(1-u-v)][1-u-\theta_i(1-u-v)]} \quad (2)$$

Thus, as shown in Figure S5 and Equation 2, if subgroup sizes are known and differences in subgroup-level seropositivity can be inferred or assumed (for example, if public health data strongly indicates differences in epidemic activity or vaccination rates between locations), allocating samples to larger subgroups and those with higher expected seropositivity will improve precision for weighted population-level seroprevalence estimates. If differences in seropositivity cannot be reasonably assumed based on available data, weighting sampling intensity by subgroup size alone can improve precision substantially (noting in Figure 1 that for any value of $\pi_1$, $\mathit{arg\,min}_{p_1} W(p_1)$ increases with increasing values of $d_1$).

**Model-based comparison between sample allocation strategies**

We first considered a system of two subgroups with total population of $N$ individuals, with $d_1$ and $d_2$ specifying the proportions of $N$ in each subgroup. We set a fixed number of total seropositive individuals, given by $N \times \theta_{\text{pop}}$ (the true seroprevalence in the entire population), and apportioned this total across the two subgroups according to the proportions $\pi_1$ and $\pi_2$. This allowed for the population seroprevalence to be fixed across different parameter combinations in each simulation. The total number of individuals that underwent serological testing is given by $n$ and the proportion of these $n$ individuals sampled from each subgroup is given by $p_1$ and $p_2$.

Procedures for the numerical simulation are as follows: (1) Generate two subgroups with sizes $d_1 \times N$ and $d_2 \times N$ and specify the true number of seropositive individuals in each subgroup as $N \times \theta_{\mathrm{pop}} \times \pi_1$ and $N \times \theta_{\mathrm{pop}} \times \pi_2$; (2) Randomly draw, without replacement, $p_1 \times n$ and $p_2 \times n$ individuals from each subgroup; (3) Incorporate serological test performance by drawing the number of observed positives from binomial distributions with probabilities equal to the test sensitivity (for true positives) or $1 -$ *specificity* (for false positives); (4) Calculate the estimated population seroprevalence via post-stratification, $\hat{\theta}_{\mathrm{pop}} = \sum_i d_i \, \hat{\theta}_i$, where $\hat{\theta}_i$ is the observed seroprevalence, adjusted for test performance, in subgroup $i$. (5) Repeat steps 1-4 10000 times to generate a distribution of $\hat{\theta}_{\mathrm{pop}}$ values. We report $W$, the width of the 95th percentile interval for each distribution, as an approximate measure of uncertainty for $\hat{\theta}_{\mathrm{pop}}$ for a given set of parameters $(d_1, d_2, \pi_1, \pi_2, p_1, p_2)$.

We extended this model to evaluate optimal sample allocation strategies in the case of heterogeneous seropositivity across multiple subgroups stratified by age and location. We used data from our Somerville, Massachusetts COVID-19 serosurveillance study and public health data on PCR testing for SARS-CoV-2 infection to inform the numerical simulation in this analysis.

To examine how choice of sampling location influence uncertainty in seroprevalence estimates obtained via convenience sampling, we used a numerical model that incorporates performance characteristics of serological testing and heterogeneity in seropositivity across geographic and age-based subgroups. (Other demographic characteristics, including race and ethnicity, are not included in this example, but the model described here can be generalized to include any number of additional characteristics relevant for stratification.)

Supplementary Figure S5 summarizes the procedures used in the extended numerical model. We assumed a simple synthetic population that is stratified over six age categories and seven geographic units (corresponding to the seven electoral wards in Somerville), informed by age distribution and ward-level population data from Somerville. To specify “true” seropositivity in each of these 42 age-location subgroups, we assumed that the cumulative number of PCR-confirmed infections reported for each ward in Somerville, divided by the population of each ward, approximates the ward-level distribution of seropositivity across Somerville (noting that SARS-CoV-2 vaccination efforts had yet to begin at the time of our study). We then assumed that the distribution of cumulative incidence across age groups matches observed distributions for reported cases in Massachusetts [], and that the age distribution of cumulative incidence is the same for each ward. We used these assumptions to populate the matrix $\mathbf{\Theta}$, where each entry $\theta_{j,k}$ is the cumulative incidence of PCR-confirmed infection for age group $j$ in ward $k$. In the subsequent analysis, we multiplied $\mathbf{\Theta}$ by different values of $m$, to adjust for the factor by which true incident cases (and true seropositive individuals) exceed detected cases, with $m$ in $\{6,7,8,\ldots,40\}$ following estimates in [4] and [5]. We assumed that $m$ is constant across locations and age groups (i.e. that case detection effort is equal across wards and age groups).

We next generated matrices describing different survey participant catchment distributions, $\mathbf{S}$, where each entry $n_{j,k}$ represents the number of serological tests performed (equal to the number of study participants) in each age-location subgroup and where $n$ represents the total number of all tests performed. We compare three different survey participant catchment distributions: (1) $\mathbf{S}_{\mathrm{op}}$, optimal allocation of sampling effort for subgroup size and underlying cumulative incidence, given by Equation (2) in the Supplementary Information; (2) $\mathbf{S}_{\mathrm{site}}$. the actual geographic distribution of participants in the Somerville venue-based sample; (3) $\mathbf{S}_{\mathrm{alt}}$, the hypothetical geographic distribution of participants at an alternative study site in Somerville Ward 1, per GPS-estimated foot traffic data at that site.

**COVID-19 serosurveillance data collected via venue-based sampling**

We use data from a venue-based convenience-sampled serosurveillance study in Somerville, Massachusetts, conducted in June 2020, as an example case illustrating how choice of study location can influence bias and uncertainty in SARS-CoV-2 seroprevalence estimates. This site for this study was adjacent to essential business that was not subject to state-mandated restrictions on in-person business activities and thus this location had a reliably high volume of visitors each day. Spanish-, Portuguese-, and English-speaking study staff wearing personal protective equipment (PPE) contacted business patrons directly as they were leaving or entering this essential business location, and individuals recruited this way were given a flyer that was used to identify "directly recruited" participants in the study. All adults over 18 years of age with no reported symptoms suggestive of active COVID-19 infection were eligible to participate in the study. Research staff reviewed study procedures (including post-study medical follow-up for patients with positive serology) and risks and benefits of study participation before offering potential participants the opportunity to provide verbal informed consent to participate.

Participants underwent testing for SARS CoV-2-specific IgG and IgM using an immunochromatographic lateral flow assay (LFA, Biomedomics, Morrisville, North Carolina, USA). Results from point-of-care antibody testing were returned to patients by a physician on the study team. In prior validation studies using confirmed clinical cases of COVID-19 as known positives and pre-pandemic discarded blood samples as known negatives, the LFA was 90% sensitive among patients tested $>$ 8 days after symptom onset and 99.2% specific [6]. The seroprevalence among all 398 participants study, adjusted for test performance but unweighted by either age or location, was 0.113; seroprevalence was 0.130 among 228 directly recruited participants.

**Other statistical analysis**

We calculated correlations between several ward-level metrics in Somerville (including *inter alia* cumulative incidence, study participants, and GPS-estimated home locations) using Pearson's correlation coefficient $r$. We evaluated the significance of these correlations via permutation testing, wherein the observed value for $r$ is compared against a null distribution of 10,000 $r$ values obtained by randomly permuting ward assignments for the observed values for each variable. $R$ code for this procedure is included at *https://github.com/tsbrown-git/COVID19serosurveillance-Somerville*.

### Supplementary Figures

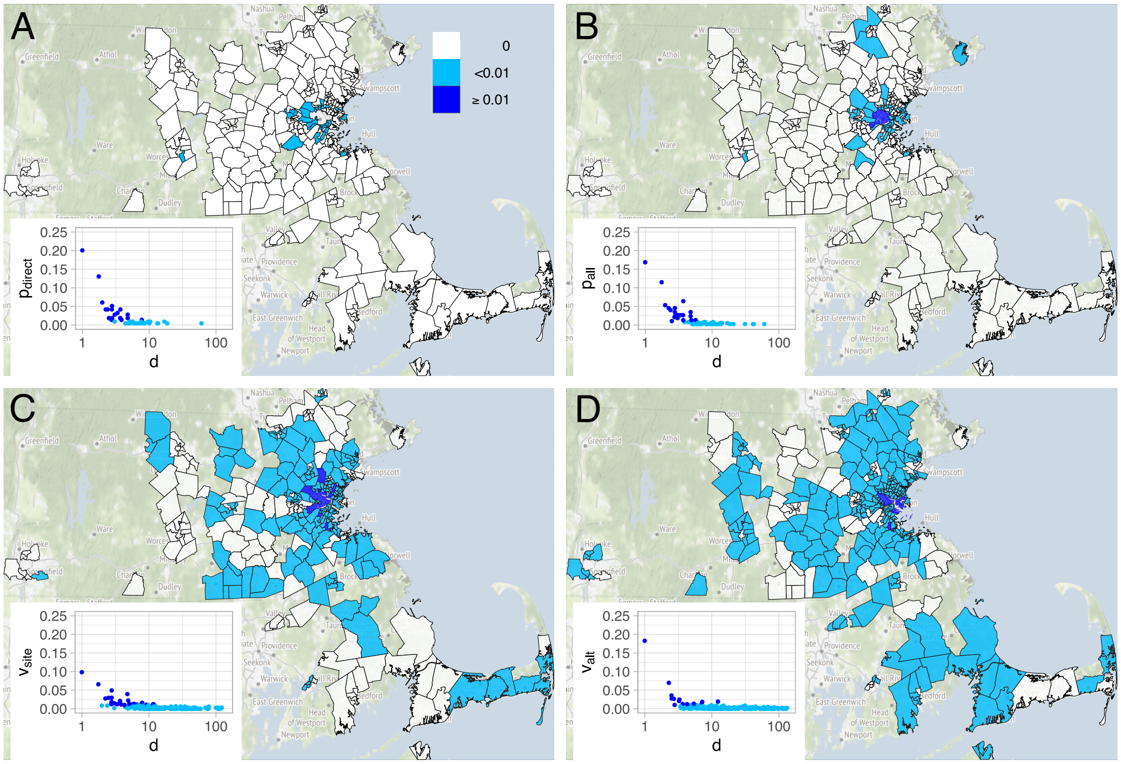


**Figure S1.** Geographic distributions of (A) self-reported reported home locations for directly-recruited participants, (B) self-reported home locations all study participants ($p_{all}$), (C) GPS-estimated visitors to the study location ($v_{site}$), and (D) GPS-estimated visitors to an alternative, hypothetical study site in Somerville Ward 1 ($v_{alt}$). Insets show proportion of participants or visitors versus distance in kilometers from the study venue. Dark map areas and points designate wards contributing > 1% of the total participants or visitors for a given site, light blue designates wards contributing ≤ 1%, and white are wards contributing zero participants or visitors.

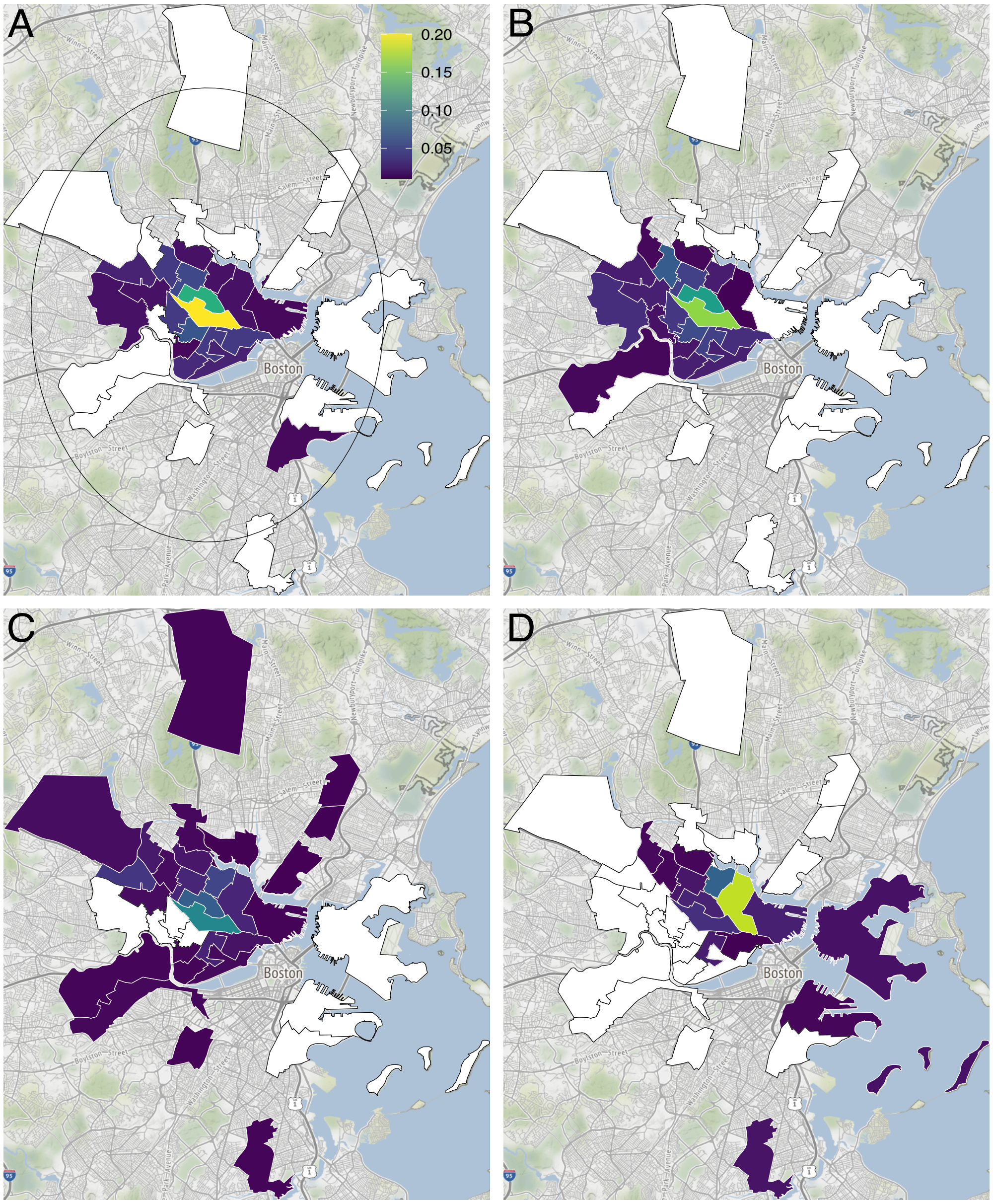


**Figure S2.** Geographic distributions of (A) self-reported reported home locations for directly-recruited participants ($p_{direct}$), (B) self-reported home locations all study participants ($p_{all}$), (C) GPS-estimated visitors to the study location ($v_{site}$), and (D) GPS-estimated visitors to an alternative, hypothetical study site in Somerville Ward 1 ($v_{alt}$), restricted to wards contributing $> 1\%$ of total number of participants or visitors. The upper right panel includes a 10 km radius circle centered on the Somerville ward containing the study venue.

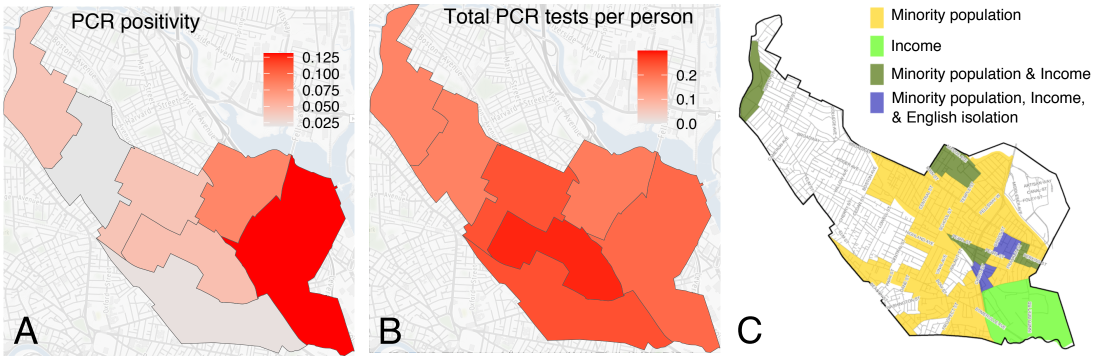


**Figure S3.** (A) Proportion of all SARS-CoV-2 test administered that were positive by Somerville electoral ward; (B) Total number of tests administered as a proportion of electoral ward population; (C) 2010 Environmental Justice Populations [7] by census block group, designated using data from the 2006-2010 American Community Survey (Minority population: >25% population identifies at non-white; Income: Households earn <65% of the statewide median household income; English isolation: >25% of households have no one over the age of 14 who speaks English only or very well.

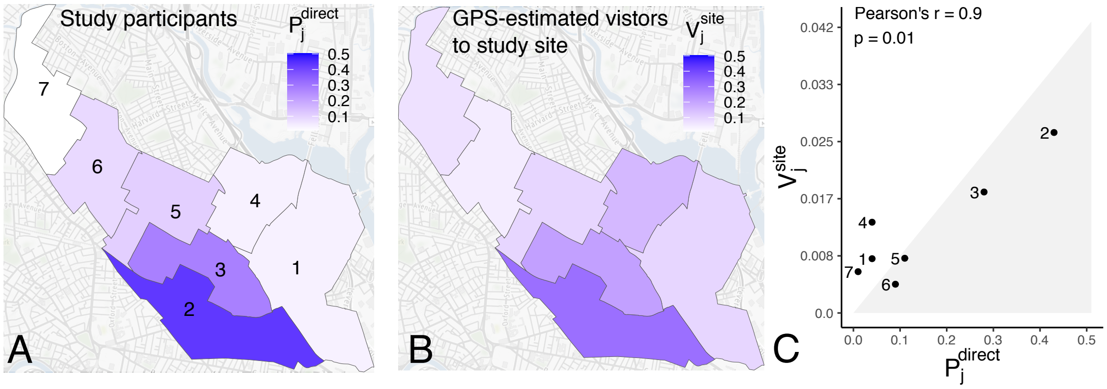


**Figure S4.** Geographic distributions of (A) proportion of all directly recruited study participants by ward ($p_{\text{direct}}$), (B) proportion of GPS-estimated visitors to the study location by ward ($v_{\text{site}}$), and (C) $p_{\text{direct}}$ versus $v_{\text{site}}$, with significance of the correlation calculated as described above.

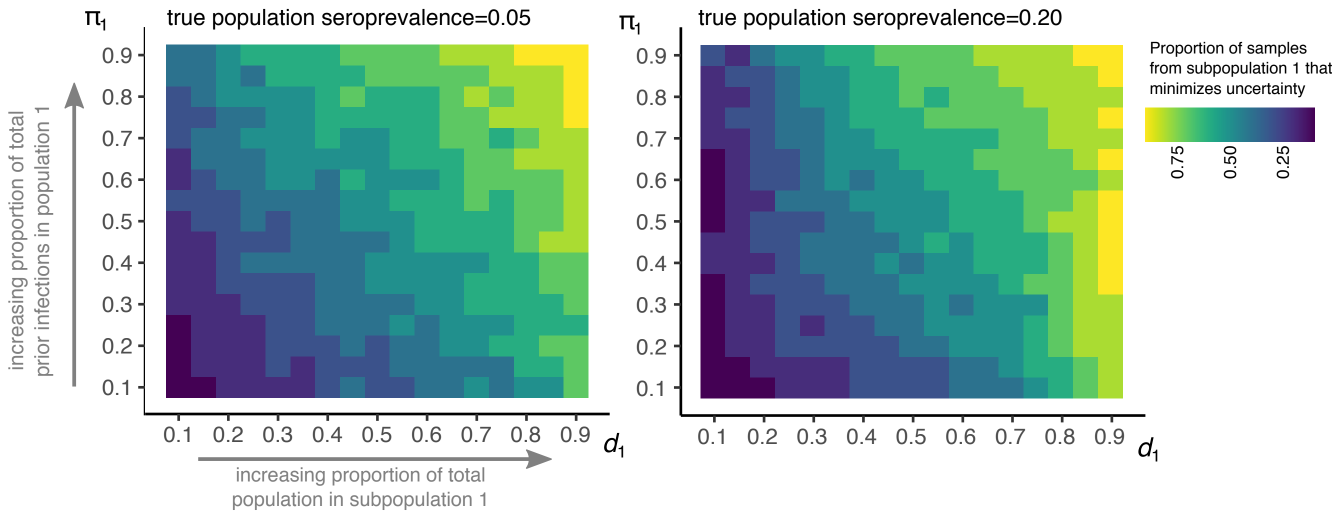


**Figure S5. Optimal sample allocation for two subgroups with varying sizes and subgroup seropositivity.** The value of $p_1$ that minimizes $W$, $arg\ min_{p_1} W(p_1)$, is estimated across multiple combinations of $d_1$ (proportion of the total population in subgroup 1) and $\pi_1$ (proportion of the total number of prior infections in subgroup 1) with true cumulative incidence $\theta_{\text{pop}} = 0.05$ or 0.2.

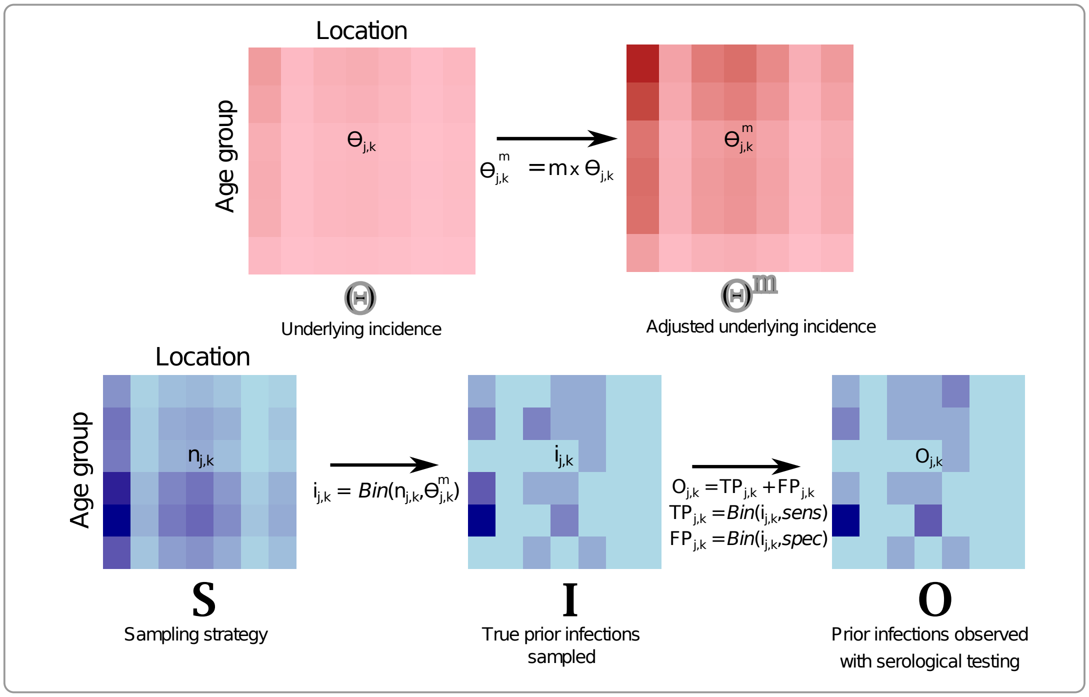


**Figure S6.** Top: Procedure for specifying true underlying seropositivity by age-location group, where entries of matrix $\boldsymbol{\Theta}$, $\theta_{j,k}$, are the estimated cumulative incidence of detected, confirmed cases (per PCR-confirmed cases reported to the City of Somerville) for individuals in location $j$ and age group $k$. Different underlying epidemic sizes are given by $\boldsymbol{\Theta^m}$, where $\theta_{j,k}^m = m \times \theta_{j,k}$ and $m$ is a multiplier approximating the factor by which true infections exceed detected, PCR-confirmed infections. Bottom: Procedures for estimating uncertainty in serology-based cumulative incidence using different sampling strategies, $\boldsymbol{S}$ (as described in Methods and Supplementary Information). The procedure in is repeated 1,000 times for each value of $m$ and $n$.

7. Massachusetts Executive Office of Energy and Environmental Affairs. Massachusetts Environmental Justice Populations. Available at: https://www.mass.gov/info-details/environmental-justice-populations-in-massachusetts
